# Comparative analysis of three point-of-care lateral flow immunoassays for detection of anti-SARS-CoV-2 antibodies: data from 100 healthcare workers in Brazil

**DOI:** 10.1101/2020.11.11.20229914

**Authors:** Danielle Dias Conte, Joseane Mayara Almeida Carvalho, Luciano Kleber de Souza Luna, Klinger Soares Faíco-Filho, Ana Helena Perosa, Nancy Bellei

## Abstract

Since the Coronavirus Disease 2019 (COVID-19) pandemic, Brazil has the third-highest number of confirmed cases and the second-highest number of recovered patients. SARS-CoV-2 detection by real-time RT-PCR is the gold standard but requires a certified laboratory infrastructure with high-cost equipment and trained personnel. However, for large-scale testing, diagnostics should be fast, cost-effective, widely available, and deployed for the community, such as serological tests based on lateral flow immunoassay (LFIA) for IgM/IgG detection. We evaluated three different commercial point-of-care (POC) LFIAs for anti-SARS-CoV-2 IgM and IgG detection in capillary whole blood of 100 healthcare workers (HCW) from São Paulo university hospital previously tested by RT-PCR: 1) COVID-19 IgG/IgM BIO (Bioclin, Brazil), 2) Diagnostic kit for IgM/IgG Antibody to Coronavirus (SARS-CoV-2) (Livzon, China); and 3) SARS-CoV-2 Antibody Test (Wondfo, China). A total of 84 positives and 16 negatives HCW were tested. The data was also analyzed by the number of days post symptoms (DPS) in three groups: <30 (n=26), 30-59 (n=42), and >59 (n=16). The observed sensibility was 85.71%, 47.62%, and 44.05% for Bioclin, Wondfo, and Livzon, respectively, with a specificity of 100% for all LFIA. Bioclin was more sensitive (*p*<0.01), regardless of the DPS. Thus, the Bioclin may be used as a POC test to monitor SARS-CoV-2 seroconversion in HCW.

## 1. Introduction

After ten months since the Coronavirus Disease 2019 (COVID-19) pandemic [1], caused by de the severe acute respiratory syndrome-related coronavirus 2 (SARS-CoV-2), Brazil reached, until late January 2021, the third place in the number of confirmed cases, accounting for more than 9.1 million cases and 222 thousand deaths [2]. However, is the second country with the highest number of recovered patients (more than 8.4 million) [3].

The molecular detection of SARS-CoV-2 by real-time Reverse Transcription-Polymerase Chain Reaction (RT-PCR) is the gold standard test but requires a certified laboratory infrastructure with high-cost equipment and trained personnel. This structure is suitable, and of paramount importance, for the diagnostic of hospitalized patients, as well as healthcare workers (HCW). However, for large-scale testing, RT-PCR is not the best option. Therefore, COVID-19 diagnostic tests should be fast, cost-effective, widely available, and deployed for the community. In general, those requisites are achieved by serological tests based on lateral flow immunoassay (LFIA). Many LFIAs have been described for the detection of IgM and IgG immunoglobulins against SARS-CoV-2, or detection of SARS-CoV-2 viral proteins (antigen tests) [4]. Many of LFIAs for SARS-CoV-2 antibody detection are manufactured using the nucleocapsid (N) or spike (S) protein, or N and S combined [5]. These tests should also be used to support RT-PCR results, especially since antibody responses can assist in the prognosis of patients [6]. Immunoglobulin response against viral infection begins with an early and transient IgM production, followed by a longer and lasting IgG response. In patients with COVID-19, the production of IgM and IgG could be simultaneous and detected after two days of symptoms onset, and could reach in some patients a plateau level after six days [7, 8]. Particularly, the IgM response could last for more than six months [9]. Moreover, the immunoglobulin levels are in most cases correlated positively with the severity of COVID-19 although the antibody response could be delayed in critical patients compared to non-critical cases [10].

In the present study, we evaluated the sensitivity of three different commercial point-of-care (POC) LFIAs for anti-SARS-CoV-2 antibody detection in 100 HCW from São Paulo university hospital in Brazil, with confirmed tests (positive or negative) for COVID-19 by real-time RT-PCR assay.

## 2. Material and methods

Three commercial POC LFIAs for detection of anti-SARS-CoV-2 IgG and IgM were tested: 1) COVID-19 IgG/IgM BIO (Bioclin, Brazil), 2) Diagnostic kit for IgM/IgG Antibody to Coronavirus (SARS-CoV-2) (Livzon, China); and 3) SARS-CoV-2 Antibody Test (Wondfo, China). Bioclin and Livzon LFIAs independently detect IgG and IgM, whereas Wondfo detects IgG and IgM combined. In brief, these tests detect IgG and IgM immunoglobulins anti-SARS-CoV-2, in a lateral flow assay, that react with colloidal gold particles conjugated with SARS-CoV-2 antibodies, which in turn are captured by antibodies against human IgM and IgG present in the Test Region (T), resulting in a dark-colored test band (positive result). A Control Region (C), present before T region, indicates a valid test when a dark-colored band is also generated, due to the reaction between human immunoglobulins and human anti-immunoglobulin antibodies fixed in the C region, or invalid otherwise. The declared combined sensitivity (IgG/IgM) of the manufacturer’s LFIAs are 96.3%, 90.6%, and 86.43% for Bioclin, Livzon, and Wondfo, respectively.

A total of 100 HCW from the São Paulo university hospital, previously tested for SARS-CoV-2 infection by real-time RT-PCR with the GeneFinder COVID-19 Plus RealAmp Kit (Osang Healthcare, Korea), in the period of March to June 2020, were enrolled in the study. From them, 84 were confirmed positive, and 16, negative.

The indicated volume of finger-prick capillary whole blood for each test, collected preferably from the skin of annular fingertip with a lancing device, was pipetted immediately into the cassette sample wells, following the addition of sample diluent according to the manufacturer instructions. All LFIAs were tested simultaneously at the moment of blood draw of each investigated HCW. Results were read up to 15 minutes to confirm negative results. The LFIAs were performed from April to July 2020.

The sensitivity was calculated as the proportion of positive results of LFIAs in relation to the positive RT-PCR confirmed cases, and specificity was calculated as the proportion of LFIAs negative results in relation to the negative RT-PCRs. The 95% confidence intervals (CI) of sensitivity and specificity proportions were calculated by the modified Wald method. The results were also analyzed according to the number of days post symptoms (DPS), distributed in three distinct groups: <30 (n=26), 30-59 (n=42), and >59 (n=16). The proportion of results accounted for IgM and IgG, alone or combined, regarding DPS, and the pairwise comparison within LIFAs was analyzed by Cochran’s Q and McNemar tests, for a *p*-value <0.05. The analysis was made using software R version 4.0.2 [11].

The study was approved by the São Paulo hospital Research Ethics Committee (CEP n. 34371020.5.0000.5505).

## 3. Results

The age of investigated HCWs varied from 20 to 67 years (mean = 37.45, median = 36). The overall sensitivity of IgM and IgG detection, individually or combined, are described in table 1.

**Table 1:** Sensitivity of LFIAs results from 84 positive RT-PCR HCW for SARS-CoV-2.

| LFIA | Sensitivity in % (95% CI) |  |  |
| --- | --- | --- | --- |
|  | IgG/IgM | IgM | IgG |
| Bioclin | 85.71 (76.52-91.79) | 54.76 (44.14-64.97) | 85.71 (76.52-91.79) |
| Livzon | 44.05 (33.92-54.70) | 29.76 (21.01-40.29) | 35.71 (26.28-46.40) |
| Wondfo | 47.62 (37.28-58.17) | N.A. | N.A. |

HCW, healthcare workers. N.A., not available.

Bioclin LFIA showed an overall sensitivity of 85.71% (72/84), followed by Wondfo with 47.62% (40/84) and Livzon with 44.05% (37/84). In comparison to the 16 negative RT-PCR individuals, the sensitivity of all LFIAs was 100% (77.31% to 100%, 95% CI).

The results showing the overlap between individual IgG and IgM reactivity for Bioclin and Livzon are shown in figure 1.

**Fig. 1.**
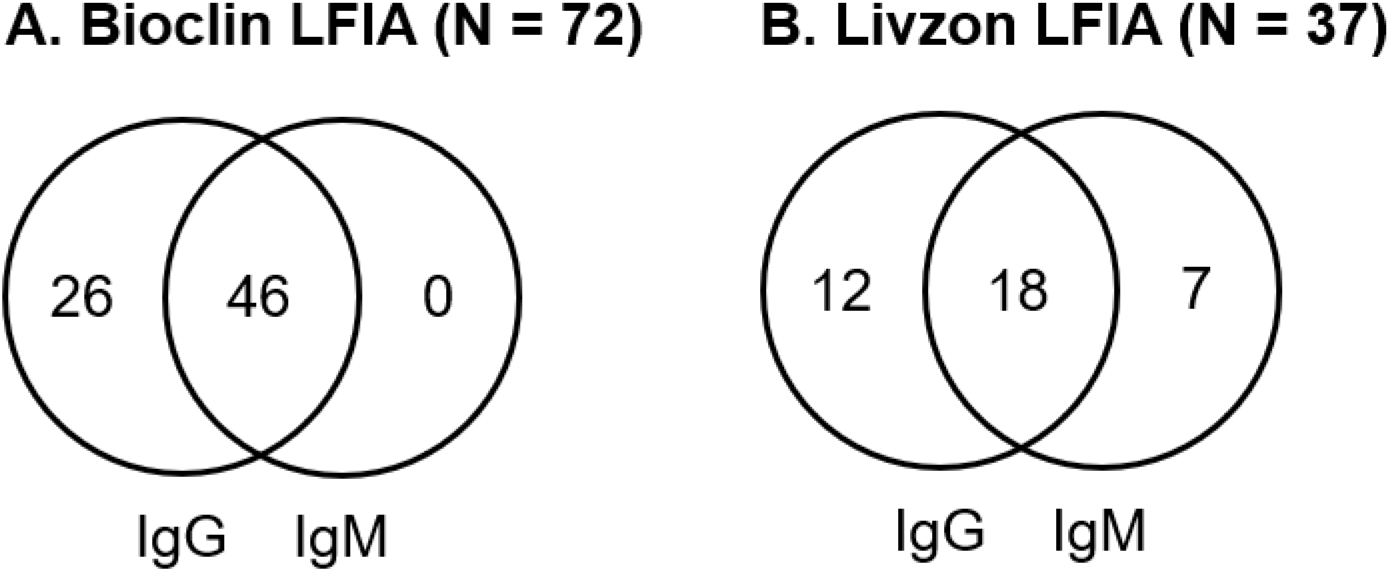
Venn diagram showing the overlap between individual IgG and IgM reactivity for Bioclin and Livzon LFIAs.

The results according to the groups of DPS (<30, 30-59, and >59), are depicted in table 2.

**Table 2:** Comparison of LFIAs results in time groups according to the days post symptoms (DPS).

| Antibody | DPS | HCW | number (%) |  |  | *p-value* |
| --- | --- | --- | --- | --- | --- | --- |
|  |  |  | Bioclin | Wondfo | Livzon |  |
| IgM/IgG<sup>1</sup> | <30 | 26 | 22 (84.62) | 14 (53.85) | 13 (50.00) | 0.0022* |
|  | 30-59 | 42 | 38 (90.48) | 21 (50.00) | 17 (40.48) | <0.001* |
|  | >59 | 16 | 12 (75.00) | 5 (31.25) | 7 (43.75) | 0.0131* |
| IgM<sup>2</sup> | <30 | 26 | 19 (73.08) | NA | 11 (42.31) | 0.0047* |
|  | 30-59 | 42 | 21 (50.00) | NA | 11 (26.19) | 0.03892* |
|  | >59 | 16 | 6 (37.50) | NA | 3 (18.75) | 0.0833 |
| IgG<sup>2</sup> | <30 | 26 | 22 (84.62) | NA | 10 (38.46) | 0.0005* |
|  | 30-59 | 42 | 38 (90.48) | NA | 15 (35.71) | <0.001* |
|  | >59 | 16 | 12 (75.00) | NA | 5 (31.25) | 0.0081 |

DPS, days post symptoms. HCW, healthcare workers.
\*significant for $p < 0.05$
<sup>1</sup>Cochran's Q test, $p < 0.05$
<sup>2</sup>McNemar test, $p < 0.05$

The Bioclin LFIA was significantly more sensitive, in comparison to Livzon and Wondfo, regardless of the DPS or detection of IgM and IgG combined (Cochran’s Q test, *p*<0.05). The posthoc analysis of pairwise comparisons, with the McNemar test, also have shown that Bioclin was more sensitive than Livzon for IgM and IgG individually, and no differences were observed between Livzon and Wondfo regardless of the DPS and immunoglobulin class (table 2).

The proportion of positive results for each LFIA test along the analyzed DPS have not shown any significant difference for the overall IgM/IgG detection (Bioclin, *p*=0.316; Livzon, *p*=0.744; Wondfo, *p*=0,33), although the sensibility of Wondfo LFIA dropped to 31.25% after 60 DPS. The same was observed for IgG (Bioclin, *p*=0.316; Livzon, *p*=0.894) and IgM (Bioclin, *p*=0.054; Livzon, *p*=0.208) alone, although Bioclin is likely to be more sensitive for IgM in the group of <30 (*p*=0.054).

We also observed in the Wondfo LFIA test a trace of red blood cells in all lateral flow test cassettes which made reading difficult in some positive results when a faint but visible T line was present.

## 4. Discussion

In the present study, we analyzed three different commercial LFIAs for the detection of anti-SARS-CoV-2 IgG and IgM in HCW. For the POC test format, capillary whole blood is more suitable than serum or plasma and does not require a laboratory infrastructure for venous blood draw and serum/plasma separation. In the three evaluated LFIAs, the recommended volume of capillary whole blood by the manufacturers is twice the volume of serum or plasma.

The use of POC based tests for rapid antibody detection can be helpful in identifying patients at different stages of infection, due to the early production of IgM followed by IgG response, although, in patients with COVID-19, the response of IgM and IgG could be simultaneous [7, 8]. Our results demonstrated that overall sensitivity achieved by Bioclin LFIA (85.71%) with whole blood samples is compared to those obtained with serum or plasma for Wondfo (from 71.7% to 85.8%) [12-14] and Livzon (86.7%) [15], in contrast to Livzon and Wondfo LFIAs which showed sensitivities below 50%.

Similar to the results here described, Santos et al. [16] have shown, for capillary whole blood, a sensitivity of 55% for the Wondfo LFIA test in HCWs, while the sensitivity in serum samples was much higher (96%). A better sensitivity for capillary whole blood with Wondfo LFIA test was reported by Silveira et al. [13] at 77.1% in 83 volunteers with positive RT-PCR results at least 10 days before the LFIA test. In a larger study with hospitalized patients, Costa et al. [12] evaluated the Wondfo LFIA, in serum samples or plasma, and obtained a sensitivity of 85.8%. In another evaluation of the Wondfo LFIA, Wu et al. [17] have shown a sensibility of 75.8% in serum samples. In a Brazilian study accessing the performance of 12 serological tests for COVID-19 diagnosis, Cota et al. [14] described an overall sensitivity for Wondfo LFIA at 71.7% in serum from symptomatic patients with confirmed SARS-CoV-2 infection. In the same manner, the Livzon LFIA, when tested in serum samples of hospitalized patients, presented a sensibility of 80% for IgM and 86.7 for IgG, with a specificity of 95% and 100% respectively.

In summary, for LFIA, antibody detection is more effective in plasma or serum samples than in whole blood, although, for POC format and large-scale testing, finger-prick capillary whole blood is more appropriate, and therefore, choosing a more sensitive test for this type of sample is of paramount importance. In this regard, Hallal et al. [18] extrapolated the sensitivity of Wondfo LFIA at 84.8%, based on pooled results of three validation studies using plasma or serum, with sensitivities varying from 81.5% to 100%, and one using whole blood (77.1%), in two nationwide surveys on the SARS-CoV-2 antibody prevalence in Brazil but using finger-prick whole blood. On that account, antibody prevalence could have been considerably underestimated.

The majority of LFIAs for SARS-CoV-2 antibody detection are manufactured using the N or S proteins, or both combined. In terms of sensitivity, tests based on N or S antigen, in general, seem to be equivalent. However, the combined use of N and S antigens have presented a better sensitivity in comparison with N and S alone [5]. In this regard, it was not possible to analyze or infer any result concerning sensitivity differences between these antigens since the manufacturers of the analyzed LFIA do not provide this information.

An advantage of the present study is due to the fact that all LFIAs were carried out simultaneously at the time of blood draw of each HCW. On the other hand, a limitation of the study was the impossibility of follow up on each HCW to observe possible variations in the detection of IgM and IgG over time, or to expand the study to include hospitalized patients or low-income individuals from the general community.

## 5. Conclusion

Bioclin LFIA demonstrated high sensitivity and specificity for IgG detection (85.71%), and a reasonable detection of IgM (54.76%), with the use of capillary whole blood in HCW. On the other hand, Livzon and Wondfo LFIAs had an overall sensitivity below 53.85% considering all analyzed conditions (DPS, and/or IgG/IgM). Thus, the Bioclin LFIA may be a suitable POC test to monitor SARS-CoV-2 seroconversion in HCW.

## Data Availability

The authors confirm that the data supporting the findings of this study are available within the article

## Acknowledgments

J.M.A.C. and L.K.S.L. are fellows of the Coordenação de Aperfeiçoamento de Pessoal de Nível Superior (CAPES), Brazil. D.D.C. is a fellow of the Conselho Nacional de Desenvolvimento Científico e Tecnológico (CNPq), Brazil. We are grateful to Anderson Scorsato for the statistical support.

## CRediT authorship contribution statement

**Danielle D. Conte:** Validation, Investigation, Data Curation, Writing - Original Draft. **Joseane M. A. Carvalho:** Investigation, Data Curation. **Luciano K. de Souza Luna:** Validation, Investigation, Formal analysis, Writing - Review & Editing, Visualization. **Klinger S. Faíco-Filho:** Investigation. **Ana H. S. Perosa:** Investigation. **Nancy Bellei:** Conceptualization, Methodology, Resources, Supervision, Writing - Review & Editing, Project administration, Funding acquisition.

